# Proliferative Arachnoiditis and Vasculitis in Central Nervous System Tuberculosis: A Retrospective Analysis of Clinical Features and Outcomes from a Tertiary Centre in India

**DOI:** 10.64898/2026.06.25.26356514

**Authors:** Abhinav Sengupta, Rinku Sarmah, Ayan Mandal, Sandeep Rao Kordcal, Arjun Kumar Agarwal, Surabhi Vyas, Arvind Kumar, Animesh Ray, Neeraj Nischal, Manish Soneja, Naveet Wig

## Abstract

**Purpose:** Central nervous system tuberculosis (CNS TB) presenting with vasculitis or arachnoiditis causes significant morbidity and mortality. The purpose of this study was to characterise the clinical spectrum and outcomes of patients diagnosed with TB arachnoiditis and TB-associated CNS vasculitis

**Methods:** A retrospective study was conducted between October 2020 and September 2023, screening patients admitted with suspected CNS TB to a tertiary care hospital. Patients diagnosed with proliferative arachnoiditis, and TB-associated CNS vasculitis were recruited. Their clinical details, follow-up records, and outcomes were assessed.

**Results:** Among 318 patients admitted with suspected CNS TB, 87 patients had complications, with follow-up data available for 69 patients. Vasculitis, spinal arachnoiditis (SA), and optochiasmatic arachnoiditis (OCA) was diagnosed in 66 (76%), 41 (47%), and 26 (30%) patients respectively. Median duration of follow-up was 490 days. Median mRS at discharge was 4. 18 (69%) OCA patients and 14 (35%) SA patients received pulse methylprednisolone. Intrathecal hyaluronidase was administered in 14 patients and thalidomide was given to 9 patients. 30 (46%) patients with vasculitis were treated with aspirin. 69 patients completed follow-up, 49% died. Among the remaining, 88.6% had improvement with treatment with a median mRS of 2 (1-3). Among patients with OCA, 3(23.1%) showed complete improvement with a median improvement of 3 points on Likert scale. In the SA patients, 19 (55.9%, 34) patients were alive on follow-up, with a median mRS of 1. Aspirin use was not associated with better mRS or survival in patients with vasculitis. A multivariable Cox proportional model showed age at diagnosis to be the only predictor of mortality (HR 1.04, 95% CI (1.01-1.08), p =0.012).

**Conclusions:** TB arachnoiditis and CNS vasculitis are severe complications of CNS TB, and management remains a challenge. The poor therapeutic response to intrathecal hyaluronidase, thalidomide, and aspirin highlights need for further larger prospective trials and search for alternative agents.

## Introduction

Tuberculous meningitis (TBM) is the most severe manifestation of central nervous system tuberculosis (CNS TB), causing mortality, morbidity and severe neurological sequelae. Although TBM accounts for only 1-2% of all TB cases and 5-10% of extrapulmonary TB, its consequences are severe, with survivors frequently experiencing disabling complications, including hydrocephalus, cognitive impairment, motor deficits, and vision loss, and leading to long-term disability and socioeconomic burden [1,2].

TBM results from hematogenous dissemination of *Mycobacterium tuberculosis*, leading to rupture of subependymal tubercles into the subarachnoid space and formation of dense basal exudates and intense basal meningeal inflammation. The subsequent inflammation causes obstructive hydrocephalus, vasculitis, and entrapment of cranial nerves and spinal nerve roots [3]. Devastating complications include optochiasmatic arachnoiditis (OCA; causing severe bilateral vision loss), spinal arachnoiditis (SA; causing irreversible radiculomyelopathy and paraplegia), and tuberculous vasculitic infarcts (leading to stroke and focal neurological deficits). These complications may be present at diagnosis or may develop paradoxically during antitubercular treatment (ATT) due to an exaggerated immune response [4].

Despite advances in microbiological diagnosis, neuroimaging, availability of potent antitubercular agents with good CNS penetration, and the use of adjunctive corticosteroids, the prognosis for patients with complicated TBM remains poor. There is limited evidence on optimal management strategies for these complications, and novel adjunctive therapies such as high-dose steroids, intrathecal hyaluronidase, thalidomide, and aspirin are under investigation but lack robust trial data [5–7].

Given the high burden and limited outcome data for these complications in Indian patients, we conducted a retrospective cohort study at a tertiary care centre to evaluate the clinical features, management, and outcomes of OCA, SA, and vasculitic infarcts in patients with TBM.

## Materials and methods

This retrospective cohort study was conducted from July 2020 to September 2023 in the Department of Medicine, AIIMS, New Delhi, to evaluate patients diagnosed with complicated CNS TB. Patients aged 14 years or older admitted to the medicine ward, ICU, or managed in the Infectious Disease (ID) clinic, who met the consensus diagnostic criteria for CNS TB, were included. Patients with incomplete data or without a minimum follow-up of three months (excluding those who expired) were excluded. IEC approval was taken vide AIIMSA4251/28.08.25.

CNS tuberculosis was classified into definite, probable, possible, and not TBM. Definite TBM required clinical signs of meningitis along with acid-fast bacilli (AFB) in cerebrospinal fluid (CSF), positive CSF culture for *Mycobacterium tuberculosis*, or a positive nucleic acid amplification test (NAAT). Probable TBM was diagnosed using clinical criteria plus a diagnostic score of ≥10 points (without imaging) or ≥12 points (with imaging). Possible TBM involved clinical criteria plus a diagnostic score of 6–9 points (without imaging) or 6–11 points (with imaging) after ruling out other diagnoses. (Supplementary)

Complicated CNS TB was identified based on clinical and imaging findings, including OCA, SA, and tubercular vasculitis (Supplementary). Data on clinical presentations, demographics, comorbidities, biochemical parameters, and CSF studies were collected. Baseline assessments included TBM Medical Research Council (MRC) staging, visual status, limb power, seizure occurrence, and consciousness levels. Outcomes were measured in terms of mortality, modified Rankin Scale (mRS) score, and subjective visual improvement on a Likert scale (0-10), which was recorded considering the retrospective nature of study. [Supplementary table 1]

The details of ATT regimens received by the patients were included: first-line ATT referred to 4-drug regimen of isoniazid, rifampicin, pyrazinamide, and ethambutol (HRZE); augmented ATT referred to addition of one or more drugs (fluoroquinolones, linezolid, or other) to the HRZE regimen; MDR regimen referred to WHO-recommended bedaquiline-based all-oral long regimen (AOLR). The use of additional interventions, namely immunomodulatory agents, intrathecal hyaluronidase, and aspirin were also recorded.

Data were managed using Microsoft Excel and analyzed with STATA version 17.0. Categorical variables were summarized as frequencies and percentages, while continuous variables were presented as mean ± standard deviation or median (range). The chi-square test, Student’s t-test, and survival analysis were applied for statistical comparisons.

## Results

318 patients were screened for TBM in the given time period. 231 patients were excluded (46 not diagnosed as TBM, and 185 patients with TBM but not complicated by OCA/SA or vasculitis). 87 patients were included in the study, with follow-up completed for 69 patients (Figure 1). The mean (SD) age was 28 (12) years and 42 (48.3%) were male. Among the included patients, 36 (41.4%) patients were diagnosed with definitive TBM, 47 (54.0%) with probable TBM, and 4 (4.6%) with possible TBM. 26 (30%) patients had OCA, 41 (47%) had SA, and 66 (76%) patients had vasculitis. The median (IQR) CCI score was 0, and 7 (8%) patients had HIV infection. 10 (11.5%) patients had a past history of TB, and 4 (4.6%) patients had a history of TB contact. The median (IQR) GCS at presentation was 11 (9-15) with a median MRC stage at presentation of 2 (2-3). The median (IQR) duration of follow-up was 490 (121-836) days (Table 1).

**Figure 1.**
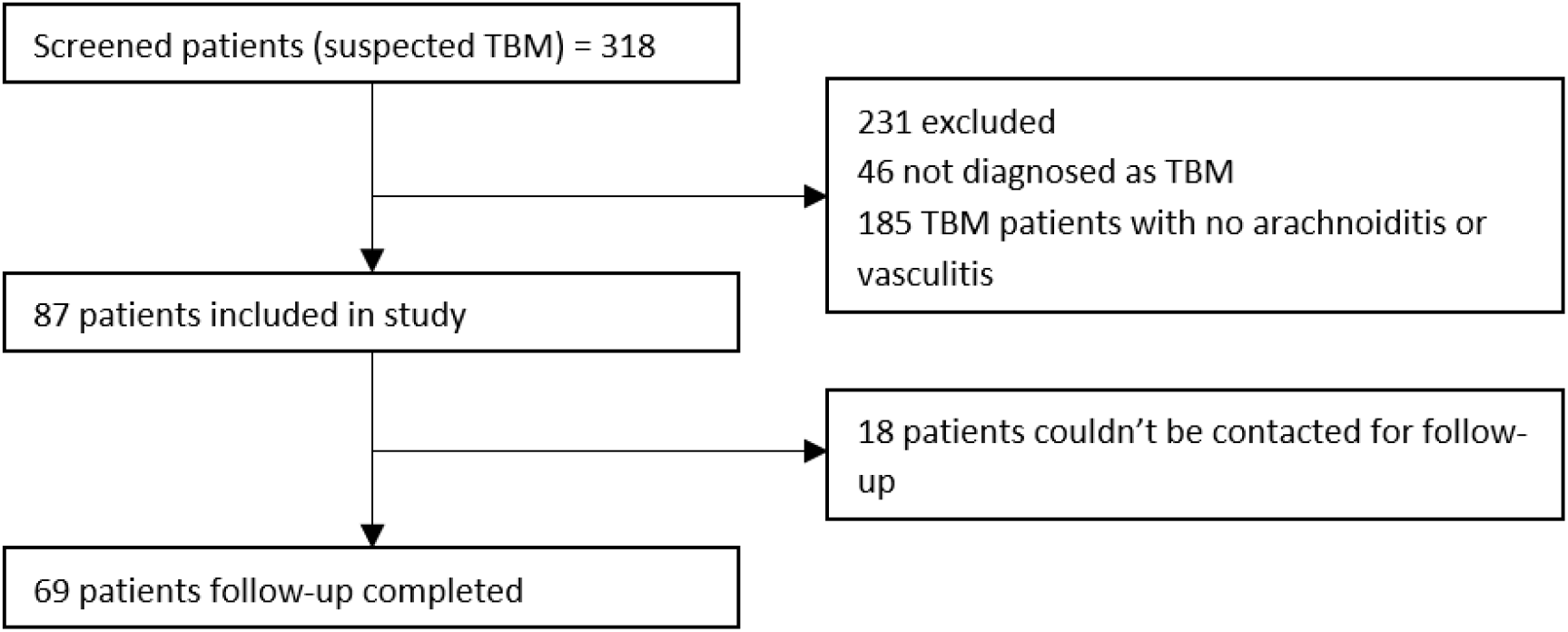
Consort diagram

**Table 1.** Baseline characteristics of patients admitted with complicated CNS TB. (n=87)

| Variable (n=87) | Result |
| --- | --- |
| Age (years) | 28.0 (12.1) |
| Sex (female) | 45 (51.7) |
| HIV positive | 7 (8.1) |
| Optochiasmatic arachnoiditis | 26 (29.9) |
| Spinal arachnoiditis | 41 (47.1) |
| Vasculitis | 66 (75.9) |
| CCI score | 0 (0- 0) |
| GCS at Presentation | 11 (9- 15) |
| MRC Stage | 2 (2- 3) |
| Hemoglobin (g/dL) | 10.8 (2.2) |
| ALC (cells/μL) | 673 (420- 1060) |
| Albumin (g/dL) | 3.5 (0.7) |
| Globulin (g/dL) | 3.3 (0.7) |
| CRP (mg/L) (n = 77) | 22.7 (7.7- 65) |
| CSF Protein (mg/dL) (n = 86) | 239.5 (115- 463) |
| CSF Sugar (mg/dL) | 34 (22- 50) |
| CSF Total Cells (/ μL) | 70 (15- 160) |
| CSF PMN (%) (n = 80) | 10 (4- 30) |
| CSF Lymphocytes (%) (n = 81) | 90 (70- 95) |
| AFB positive in CSF | 3 (3.5) |
| GeneXpert Positive | 30 (34.5) |
| Need for Mechanical Ventilation | 38 (43.7) |
| First-line ATT | 56 (64.8) |
| Augmented ATT | 24 (27.6) |
| MDR ATT | 7 (8.1) |
| Aspirin given | 32 (36.8) |
| Methylprednisolone given | 23 (26.4) |
| Hyaluronidase given | 15 (17.2) |
| Thalidomide given | 9 (10.3) |
| mRS at Discharge (n=70) | 5 (4- 5) |

**Table 2.**
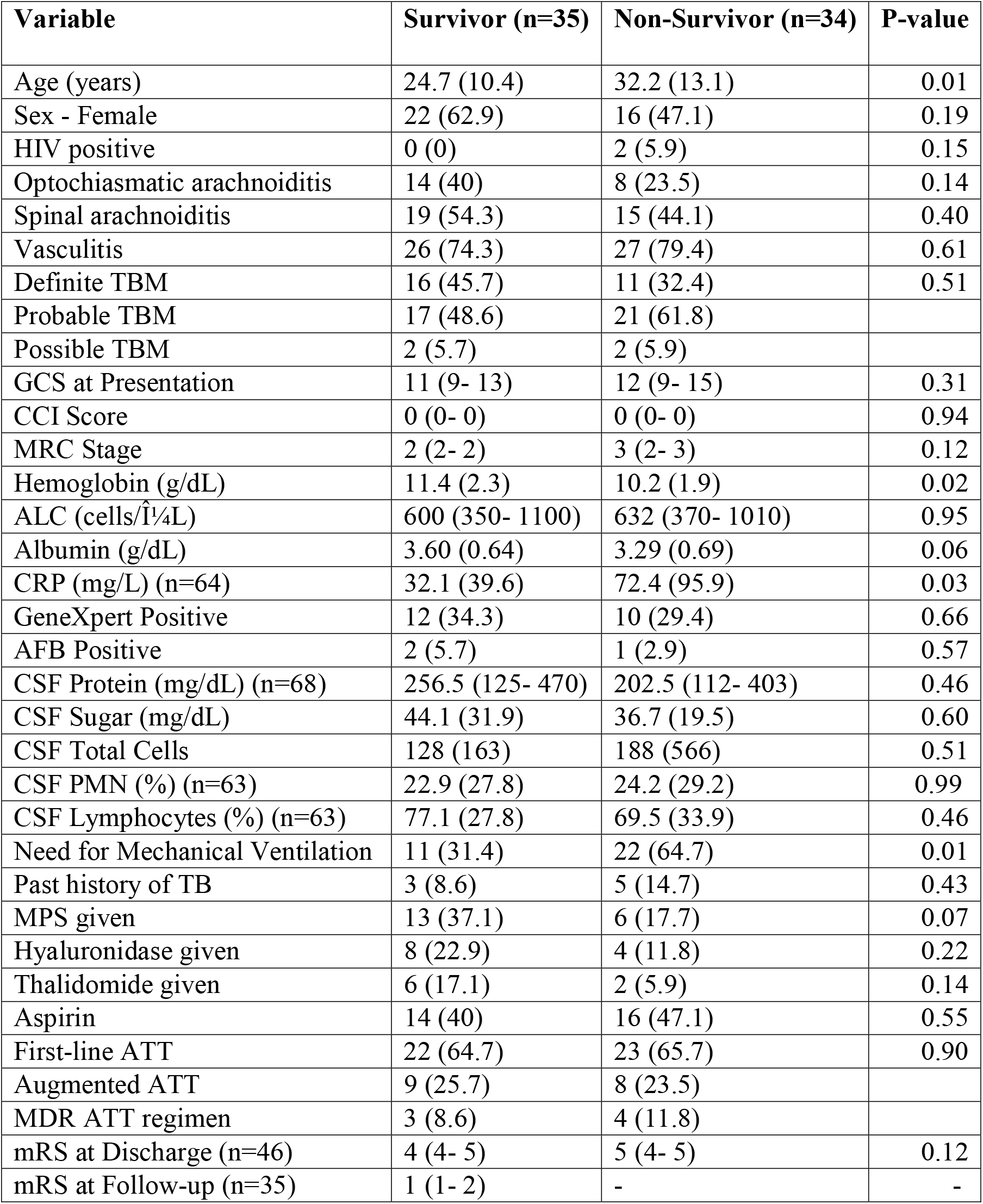
Comparison of clinico-demographic characteristics among survivors and non-survivors. (n=69)

| Variable | Survivor (n=35) | Non-Survivor (n=34) | P-value |
| --- | --- | --- | --- |
| Age (years) | 24.7 (10.4) | 32.2 (13.1) | 0.01 |
| Sex - Female | 22 (62.9) | 16 (47.1) | 0.19 |
| HIV positive | 0 (0) | 2 (5.9) | 0.15 |
| Optochiasmatic arachnoiditis | 14 (40) | 8 (23.5) | 0.14 |
| Spinal arachnoiditis | 19 (54.3) | 15 (44.1) | 0.40 |
| Vasculitis | 26 (74.3) | 27 (79.4) | 0.61 |
| Definite TBM | 16 (45.7) | 11 (32.4) | 0.51 |
| Probable TBM | 17 (48.6) | 21 (61.8) |  |
| Possible TBM | 2 (5.7) | 2 (5.9) |  |
| GCS at Presentation | 11 (9- 13) | 12 (9- 15) | 0.31 |
| CCI Score | 0 (0- 0) | 0 (0- 0) | 0.94 |
| MRC Stage | 2 (2- 2) | 3 (2- 3) | 0.12 |
| Hemoglobin (g/dL) | 11.4 (2.3) | 10.2 (1.9) | 0.02 |
| ALC (cells/$\hat{I}$¼L) | 600 (350- 1100) | 632 (370- 1010) | 0.95 |
| Albumin (g/dL) | 3.60 (0.64) | 3.29 (0.69) | 0.06 |
| CRP (mg/L) (n=64) | 32.1 (39.6) | 72.4 (95.9) | 0.03 |
| GeneXpert Positive | 12 (34.3) | 10 (29.4) | 0.66 |
| AFB Positive | 2 (5.7) | 1 (2.9) | 0.57 |
| CSF Protein (mg/dL) (n=68) | 256.5 (125- 470) | 202.5 (112- 403) | 0.46 |
| CSF Sugar (mg/dL) | 44.1 (31.9) | 36.7 (19.5) | 0.60 |
| CSF Total Cells | 128 (163) | 188 (566) | 0.51 |
| CSF PMN (%) (n=63) | 22.9 (27.8) | 24.2 (29.2) | 0.99 |
| CSF Lymphocytes (%) (n=63) | 77.1 (27.8) | 69.5 (33.9) | 0.46 |
| Need for Mechanical Ventilation | 11 (31.4) | 22 (64.7) | 0.01 |
| Past history of TB | 3 (8.6) | 5 (14.7) | 0.43 |
| MPS given | 13 (37.1) | 6 (17.7) | 0.07 |
| Hyaluronidase given | 8 (22.9) | 4 (11.8) | 0.22 |
| Thalidomide given | 6 (17.1) | 2 (5.9) | 0.14 |
| Aspirin | 14 (40) | 16 (47.1) | 0.55 |
| First-line ATT | 22 (64.7) | 23 (65.7) | 0.90 |
| Augmented ATT | 9 (25.7) | 8 (23.5) |  |
| MDR ATT regimen | 3 (8.6) | 4 (11.8) |  |
| mRS at Discharge (n=46) | 4 (4- 5) | 5 (4- 5) | 0.12 |
| mRS at Follow-up (n=35) | 1 (1- 2) | - | - |

On neuroimaging, 47 (54.0%) patients had hydrocephalus, 59 (67.8%) had tuberculoma, 61 (70.1%) had one or more vasculitic infarcts, 27 (31.0%) had perichiasmal enhancement, and 14 (16.1%) had optic nerve enhancement. Systemic imaging showed signs of dissemination in 79.1% (n/N= 68/86) of patients. The median (IQR) cells in CSF were 70 (15-160); 8 had nil cells in CSF. The median (IQR) CSF protein was 239.5 mg/dl (115-463) and CSF sugar was 34 mg/dl (22-50). On microbiological analysis, AFB was seen in 3.5 %, 34.5% patients had a positive NAAT, and CSF liquid culture was positive in 17.0% (n/N= 9/53) patients. Extra neural microbiologically proven TB was observed in 20.7% (18) patients.

56 (64.4%) patients received first line ATT, 24 (27.6%) received augmented ATT and 8 (9.2%) patients were treated with MDR regimen. Drug induced liver injury was seen in 35.6% patients. All patients received steroids as per CNS TB steroid protocol. For management of complications, pulsed methylprednisolone was given to 23 (26.4%) patients, and intrathecal hyaluronidase was administered to 15 (17.2%) patients. 9 (10.3%) patients received thalidomide, and 32 (36.8%) patients received aspirin. Neurosurgical intervention was required in 32 (37.2%), either as ventriculoperitoneal (VP) shunt (29 patients) or external ventricular drainage (EVD) (3 patients).

Follow-up data was complete in 69 patients. 34 (49%) patients expired; 12 (17.4%) died during hospital stay and 22 (31.9%) died after discharge. Among the patients who were discharged, the median (IQR) mRS at discharge was 5 (4-5). Among 35 long-term survivors, 31 (88.6%) showed improvement with treatment, median (IQR) mRS improvement being 2 (1-3).

In the OCA cohort, 18 (69.2%) patients received pulsed steroids, 11 (42.3%) received intrathecal hyaluronidase, and 9 (34.6%) were given thalidomide. Visual follow-up was available in 15 of the 26 patients. 4 (26.7%) showed complete improvement in vision, 9 (60%) had partial improvement, and 2 (13.3%) had no visual improvement. Median (IQR) improvement in vision was 3 (2-10). None of the treatments (pulsed methylprednisolone (p=0.48), intrathecal hyaluronidase (p=0.43), and thalidomide (p=0.24)) were significantly associated with a 50% or more improvement in subjective visual score (Table 3).

**Table 3.** Visual improvement with specific interventions.

| Intervention | Vision improvement | Yes | No | p-value |
| --- | --- | --- | --- | --- |
| Methylprednisolone pulse (n= 15) | Yes | 3 | 7 | 0.264 |
|  | No | 3 | 2 |  |
| Thalidomide (n= 15) | Yes | 2 | 3 | 1 |
|  | No | 4 | 6 |  |
| IT hyaluronidase (n= 14) | Yes | 2 | 3 | 1 |
|  | No | 3 | 6 |  |

Among 41 patients with SA, 19 patients were alive at follow up, with a median (IQR) mRS at follow-up of 1 (1-2), and improvement of 2 (1-3). 14 (34.2%) patients received pulse steroids; intrathecal hyaluronidase was used in 12 (29.3%) and thalidomide was used in 6 (14.6%) patients. There was no difference in mRS improvement scores among patients who received MPS pulse (p=1.0), IT hyaluronidase (p=0.64), or thalidomide (p=0.77). Follow-up was completed 34 patients with SA; mortality was observed in 15 (44.1%) patients. 8 (23.5%) of 34 patients were asymptomatic with imaging-proven SA, and all were alive at follow-up with a median mRS score of 1.5 (Table 4). In addition, 13 (14.9%) patients had coexisting OCA and SA at diagnosis.

**Table 4.** Comparison of clinico-demographic parameters and outcomes among patients with clinically manifest vs imaging-only spinal arachnoiditis.

| Variable | Clinical spinal arachnoiditis (n=26) | Imaging only spinal arachnoiditis (n=8) | p-value |
| --- | --- | --- | --- |
| Age (years) | 26.6 (8.6) | 29.5 (13.8) | 0.48 |
| OCA Present | 11 (42.3%) | 1 (12.5%) | 0.12 |
| Vasculitis Present | 19 (73.1%) | 3 (37.5%) | 0.07 |
| GCS at presentation | 11.4 (3.4) | 13.1 (2.9) | 0.17 |
| MRC Stage | 2 (2- 3) | 1 (1- 2) | 0.02 |
| CSF Total Cell Count | 237.8 (650.3) | 197.5 (184.4) | 0.05 |
| CSF Lymphocytes (%) (n=31) | 79.2 (30.6) | 81.5 (9.8) | 0.26 |
| CSF Sugar (mg/dL) | 35.2 (18.8) | 42.1 (25.4) | 0.40 |
| CSF Protein (mg/dL) (n=33) | 545.8 (1031.5) | 587.5 (605.0) | 0.15 |
| Aspirin used | 18 (69.2%) | 1 (12.5%) | 0.01 |
| Methylprednisolone pulse given | 10 (38.5%) | 2 (25.0%) | 0.49 |
| Hyaluronidase given | 10 (38.5%) | 1 (12.5%) | 0.17 |
| Thalidomide used | 5 (19.2%) | 1 (12.5%) | 0.66 |
| mRS at Discharge (n=27) | 5 (4- 5) | 4 (3- 4) | 0.01 |
| mRS at Follow-up | 6 (2- 6) | 1 (1- 1.5) | 0.00 |
| Death | 15 (57.7%) | 0 (0%) | 0.00 |

66 patients had vasculitic infarcts, and follow-up was available for 53 patients. Among the patients who survived, the median (IQR) improvement in mRS score was 3 (2-3). 30 (45.5%) patients were treated with aspirin, with no difference in mRS improvement (p=0.374), and mortality (p=0.685) among those treated. 27 patients expired on follow-up. On survival analysis, there was no difference among patients who had received aspirin for vasculitis. (HR=1.37, p=0.454) (Figure 2).

**Figure 2.**
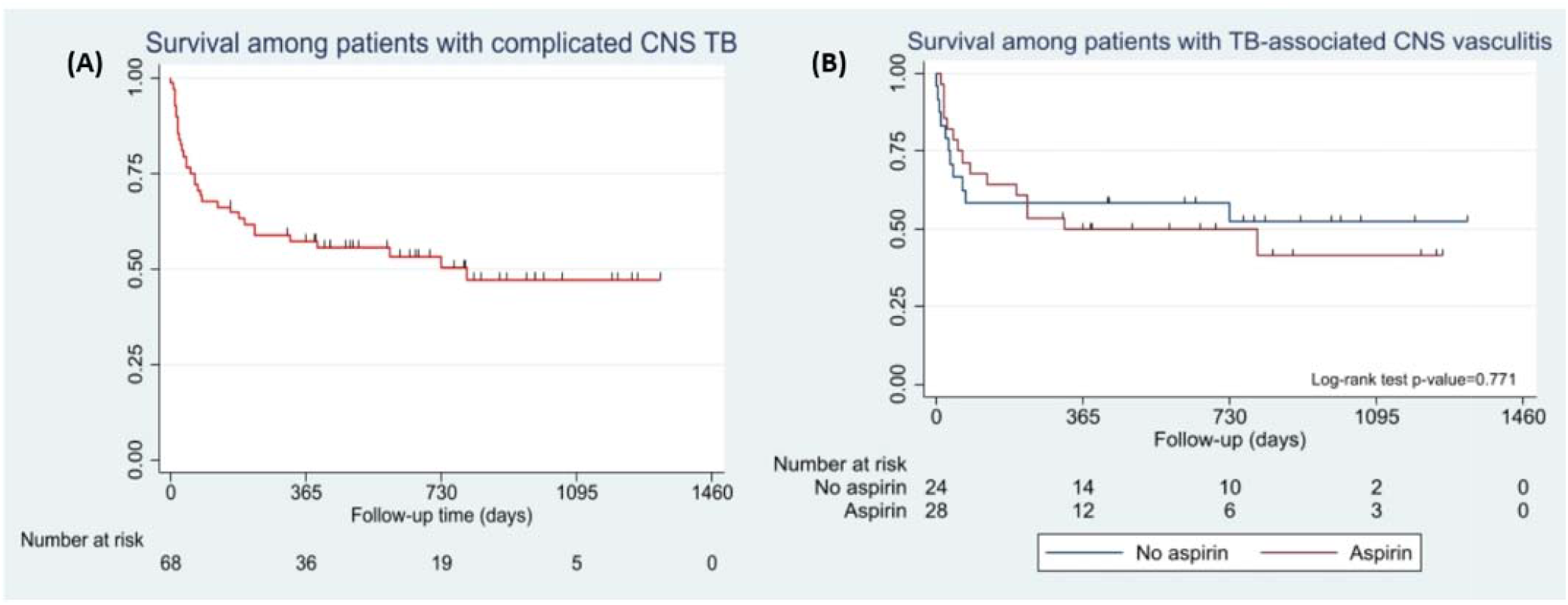
Survival analysis: (A) K-M curve showing survival among patients with complicated CNS TB (B) K-M curve analysis comparing the outcome among patients with TB-associated CNS vasculitis receiving and not receiving aspirin.

The univariable Cox proportional hazards model showed that age at diagnosis, hemoglobin, and CRP at presentation were significantly associated with mortality. A multivariable model was constructed with all parameters with a p-value <0.1, which revealed age at diagnosis to be the only independent predictor of mortality (HR 1.04, 95% CI (1.01-1.08), p =0.012).

## Discussion

CNS involvement in TB remains one of the most severe forms of the disease, with significant morbidity and mortality. OCA, SA, and vasculitic infarcts are increasingly recognized and contribute substantially to poor clinical outcomes. In our cohort of 185 patients diagnosed with TBM, such complications were identified in 87 patients (27.3%), consistent with existing literature[4,8].

OCA typically manifests as a subacute, progressive, usually bilateral near-complete loss of vision. Basal exudates containing caseous necrotic material envelop the optic chiasm, disrupting blood supply and leading to optic nerve ischemia and atrophy. OCA is reported at presentation in 10-20% of TBM cases, although it commonly develops as a paradoxical reaction after initiating ATT, which triggers intense inflammation and degeneration [9,10]. Optochiasmatic tuberculomas can coexist with arachnoiditis and may similarly appear paradoxically. Together, these lesions have been reported in up to 18% of TBM patients at diagnosis in some series [4]. Visual deterioration usually occurs within 2-3 months of starting ATT, but can manifest between 2 weeks to 18 months and can be confused with treatment failure[4]. In our study, only 26.7% of patients with OCA achieved complete visual recovery. Use of pulsed methylprednisolone, intrathecal hyaluronidase, and thalidomide did not show statistically significant associations with visual improvement.

Spinal involvement in tuberculosis is relatively common but frequently under-reported. It can present at TBM diagnosis or may develop paradoxically during ATT initiation[11]. Gupta et al. reported spinal involvement in 31% of TBM patients at presentation, with an additional 15% developing paradoxical manifestations [12]. Garg et al. reported an overall incidence of 39% of radiculomyelopathy and arachnoiditis associated with TBM [13]. In our study, SA was identified in 47% of patients, consistent with prior reports. Importantly, asymptomatic spinal involvement has been documented; such patients may develop new-onset symptoms weeks to months after starting ATT, raising concerns of treatment failure or emerging drug resistance. Srivastava et al. reported an incidence of 18.7% of asymptomatic spinal arachnoiditis TBM patients, diagnosed radiologically [14]. 23.5% patients in our study cohort had asymptomatic spinal involvement on imaging, although none of them developed new onset SA on follow up. Outcomes were poor in patients who had preexisting symptomatic SA at diagnosis, with a mortality rate of 44.1% during follow-up and only modest functional recovery in survivors.

Vasculitic infarcts can result from intense basal exudates causing obliterative arteritis, particularly affecting the Circle of Willis and perforator arteries. Reported incidence rates of stroke in TBM vary widely; 15–57% of patients develop infarctions during the course of illness [15]. Infarcts usually occur early in disease course, presenting at diagnosis or developing within the first few weeks of therapy, when inflammation is at its peak. Paradoxical new infarcts are relatively less common than other paradoxical lesions, but may occur if hydrocephalus acutely worsens or a new vasculitic focus develops. In a study by Dian et al, 60% of TBM patients had acute infarcts on baseline neuroimaging, while only 1 patient showed a new infarct on follow-up MRI(16). Risk of stroke strongly correlates with disease severity, with most of them occurring in advanced disease (stage II–III) where basal exudation and vasculitis are pronounced [15]. In our study, the incidence of vasculitic infarcts was 75.9%, higher than reported in previous literature. Although the median improvement in mRS in our patients on follow up was 3, mortality was high (40.9%), likely reflecting the inclusion of sicker patients requiring hospital admission.

A substantial proportion of patients in our cohort had evidence of TB dissemination (79.1%). Disseminated disease is often associated with a high mycobacterial load and an impaired immune response. Both these factors contribute to disease spread and also have a potential to develop an exaggerated inflammatory response on treatment initiation. This is likely due to abrupt immune engagement with previously unrecognized or poorly contained antigens[17,18].

Timely recognition and management of complications is crucial to improve functional outcomes. Mainstay of treatment is continuation of ATT along with appropriate immunomodulation. Although standardized treatment protocols are lacking, several agents such as high-dose pulse methylprednisolone, dexamethasone, and thalidomide have been used with variable success[6,7,9,10,19]. Intrathecal hyaluronidase has been used in select cases to break down the dense fibrinous adhesions in SA, although supportive evidence remains limited to older case series [20,21]. Neurosurgical management (VP shunting and EVD) may be considered in patients with hydrocephalus refractory to medical therapy [9,10,22]. Despite aggressive therapy, neurological recovery remains unsatisfactory with high mortality [23]. Aspirin has been trialled as an adjunctive therapy to reduce infarct burden and improve outcomes. While a few studies have suggested potential benefits of aspirin [5,24], robust evidence from larger trials remains limited, particularly in patients with advanced or disseminated disease. In our study, the use of aspirin was not associated with a statistically significant reduction in mortality or improvement in mRS scores.

This study has several limitations. First, the retrospective nature of the study introduces the risk of selection and reporting bias, limiting causality establishment. Second, sicker admitted patients were recruited, which might overestimate the true incidence of complications associated with TBM. Third, although the cohort is relatively large for a single-center study of TBM with paradoxical manifestations, heterogeneity in treatment protocols (especially the use of steroids, thalidomide, hyaluronidase, and aspirin) limits ability to draw definitive conclusions regarding treatment efficacy. These interventions were administered based on clinician discretion rather than standardized criteria. Fourth, follow-up data were incomplete for a subset of patients, potentially underestimating long-term outcomes and mortality. Finally, the use of subjective visual scales and mRS scores may not fully capture the spectrum of neurological impairment, particularly in patients with subtle cognitive or sensory deficits.

OCA, SA and vasculitis represent severe and increasingly recognized complications of TBM that are associated with high morbidity and mortality. Despite timely initiation of ATT and adjunctive immunosuppressive interventions, outcomes remain poor. Our findings highlight the importance of early identification, individualized management, and long-term follow-up in this high-risk cohort. Larger prospective multicenter studies are required to establish standardized treatment protocols and evaluate the efficacy of adjunctive therapies to improve long-term neurological and functional outcomes.

## Supporting information

STROBE Checklist

## Data Availability

All data produced in the present study are available upon reasonable request to the authors

## Author contributions

Conceptualization: Abhinav Sengupta, Manish Soneja

Methodology: Manish Soneja, Abhinav Sengupta

Formal Analysis: Abhinav Sengupta

Investigation (Data Collection): Abhinav Sengupta, Rinku Sarmah, Ayan Mandal, Sandeep Rao Kordcal, Arjun Kumar Agarwal, Arvind Kumar, Neeraj Nischal, Surabhi Vyas

Data Curation: Abhinav Sengupta

Writing – Original Draft: Abhinav Sengupta, Ayan Mandal, Sandeep Rao Kordcal

Writing – Review & Editing: Ayan Mandal, Animesh Ray, Manish Soneja, Naveet Wig

Visualization: Abhinav Sengupta

Supervision: Manish Soneja, Animesh Ray, Naveet Wig

Project Administration: Manish Soneja, Ayan Mandal

## Acknowledgements

We sincerely thank the patients and their relatives who participated in the study and dedicated their valuable time to sharing their experiences, without whom this study would not have been possible.

## Access to data

De-identified study data can be made available on reasonable request.

