## Supplementary for "Proliferative Arachnoiditis and Vasculitis in Central Nervous System Tuberculosis: A Retrospective Analysis of Clinical Features and Outcomes from a Tertiary Centre in India"

**Definitions:**

Consensus definition for diagnosis of TBM

Clinical entry criteria

• Symptoms and signs of meningitis including one or more of the following: headache, irritability, vomiting, fever, neck stiffness, convulsions, focal neurological deficits, altered consciousness, or lethargy.

Tuberculous meningitis classification

Definite tuberculous meningitis

• Patients should fulfill criterion A or B:

A)	Clinical entry criteria plus one or more of the following: acid-fast bacilli seen in the CSF; Mycobacterium tuberculosis cultured from the CSF; or a CSF positive commercial nucleic acid amplification test.

B)	Acid-fast bacilli seen in the context of histological changes consistent with tuberculosis in the brain or spinal cord with suggestive symptoms or signs and CSF changes, or visible meningitis (on autopsy).

Probable tuberculous meningitis

• Clinical entry criteria plus a total diagnostic score of 10 or more points (when cerebral imaging is not available) or 12 or more points (when cerebral imaging is available) plus exclusion of alternative diagnoses. At least 2 points should either come from CSF or cerebral imaging criteria.

Possible tuberculous meningitis

• Clinical entry criteria plus a total diagnostic score of 6–9 points (when cerebral imaging is not available) or 6–11 points (when cerebral imaging is available) plus exclusion of alternative diagnoses. Possible tuberculosis cannot be diagnosed or excluded without doing a lumbar puncture or cerebral imaging.

Not tuberculous meningitis

• Alternative diagnosis established, without a definitive diagnosis of tuberculous meningitis or other convincing signs of dual disease.

Complicated CNS tuberculosis will be diagnosed as per the following criteria:

Optochiasmatic arachnoiditis Vision loss with imaging evidence of arachnoiditis- optochiasmatic arachnoiditis/optic nerve arachnoiditis/ dense basal exudates

Spinal arachnoiditis: Paraparesis/quadriparesis/sphincter dysfunction due to spinal radiculomyelitis with imaging evidence of spinal arachnoiditis.

Tubercular vasculitis: Imaging suggestive of acute infarction (T2/FLAIR hyperintense with diffusion restriction) or imaging evidence of vascular involvement (vessel narrowing/occlusion with perivascular enhancement) with or without clinical focal neurological deficits

Modified Rankin scale

| **Grade** | **mRS** |
| --- | --- |
| 0 | No symptoms at all |
| 1 | No significant disability: despite symptoms, able to carry out all usual duties and activities |
| 2 | Slight disability: unable to perform all previous activities but able to look after own affairs without assistance |
| 3 | Moderate disability: requiring some help but able to walk without assistance |
| 4 | Moderately severe disability: unable to walk without assistance and unable to attend to own bodily needs without assistance |
| 5 | Severe disability: bedridden, incontinent and requiring constant nursing care and attention |
| 6 | Death^*^ |
